# Modeling geographical spread of COVID-19 in India using network-based approach

**DOI:** 10.1101/2020.04.23.20076489

**Authors:** Ankush Kumar

## Abstract

COVID-19 pandemic is a global concern, due to its high spreading and alarming fatality rate. Mathematical models can play a decisive role in mitigating the spread and predicting the growth of the epidemic. India is a large country, with a highly variable inter-state mobility, and dynamically varying infection cases in different locations; thus, the existing models, based solely on the aspects of growth rates, or generalized network concepts, may not provide desired predictions. The internal mobility of a country must be considered, for accurate prediction. Herein, we propose a framework for predicting the geographical spread of COVID-19, using reported COVID-19 cases, census migration data, and monthly airline data of passengers. We discover that spreading depends on the spatial distribution of existing cases, human mobility patterns, and administrative decisions. In India, the mobility towards professional sites can surge incoming cases at Maharastra and Karnataka, while migration towards the native places can risk Uttar Pradesh and Bihar. We anticipate that the state Kerala, with one of the highest cases of COVID-19, may not receive significant incoming cases, while Karnataka and Haryana may receive the challenge of high incoming cases, with medium cases so far. Using airline passenger’s data, we also estimate the number of potential incoming cases at various airports. The study predicts that the airports located in the region of north India are vulnerable, whereas in northeast India and in some south India are relatively safe. The detailed analysis in this direction will guide policymakers for prior planning of transport, and minimize the spread of COVID-19.

## I. INTRODUCTION

COVID-19 is an actively spreading pandemic in the whole globe and is an unprecedented challenge for the healthcare, economy, and lifestyle of the community.[1–3] Countries are striving hard to mitigate the spread of COVID-19 by various strategies: banning gathering, closing schools, stopping transports, locking down cities, imposing curfews, and sealing locations, and still not able to effectively contain it. The need of the hour is to get location by location risk assessment so that timely preventive measures can be taken. Researchers have systematically studied various aspects related to COVID-19, such as the role of isolation of cases and their contacts,[4] impact of non-pharmaceutical interventions,[5] obtaining infected population from the death counts,[6, 7],and calculating optimum duration [8] and effectiveness of lock-down period[5, 9, 10]. The available research in this area is primarily on analyzing growth in the number of infectious cases in the local community.[8, 11, 12] These models mainly use non-linear fittings on time series of reported cases in a particular region to estimate the time evolution of epidemics in that region.

Human mobility and transport also play critical roles in the spread of COVID-19, adding seeds of disease-transmission. However, there is a limited effort in the literature to model the impact of human mobility on the spread of COVID-19, particularly within a country. Chinazzi et al. and Wells et al. studied the importance of the travel ban of China and important border policies.[13, 14] Paster et al. used a Long short-term memory (LSTM) based neural network to predict the risk category of a country. Pujari et al. attempted multicity model and assumed the fraction of population reaching a neighbor is inversely proportional to its degree connections.[15]

COVID-19 currently affects almost all developed and developing countries; India is one of the countries with significant COVID-19 cases. India is currently in stage 2 of epidemics, and strict plans and steps are required to prevent it from entering stage 3 or higher. India is a large country, with diverse cultures, languages, jobs, and educational opportunities, resulting into distinct and complex connection patterns between different locations. Thus, a generic mathematical network approach [16–18] such as a small world and scale-free model can not be employed for disease-spread analyzing for India; important geographical aspects of human-mobility should also be incorporated in a model. To tackle it, herein, we propose a network-based framework for modeling COVID-19 risk at different geographical locations by using migration and airflow based real data. The proposed model can be used for policymaking, regulating transport, and predicting future hotspots.

## II. RESULTS AND DISCUSSION

The model consists of dividing the space (say country) into various components, which can be states, districts or cities depending on the available data. Consider there are N components with the population of individual components *P*_*i*_ and infected numbers *I*_*i*_. The probability, *p*_*i*_ of an infected person in the population *P*_*i*_ can be written as

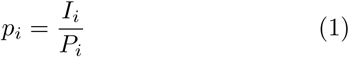

Now, let, *T*_*ij*_ is the number of transported individuals from *i* to *j* component, then, a certain number of infected individuals would also be transferred from location *i* to *j* as follows:

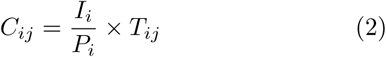

Thus, the total number of incoming cases at destination (*j*) from all connected components(*i*) can be written as

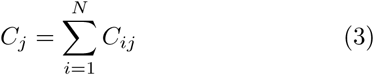

To model COVID-19 spread in India, here we use the states and union territories as components and their reported positive cases as the number of people infected. Mobility between two states relies on the intermixed community; migration may be a strong indicator of this. Thus, to analyze the potential inter-state mobility, interstate migration data is obtained from the census 2011.[19]To obtain native to professional (work or education) place-based migration, only the data of male entries are considered to avoid significant marriage-related migration. Figure 1(a) represents the migration map between different states with arrow-profile; here, the arrow width represents the migration number and the arrow direction is from native to the professional place. As seen from the arrow widths, the migration nature is unsymmetrical with high variability. The highest migration takes place between Uttar Pradesh to Maharashtra, Uttar Pradesh to Delhi, Bihar to Delhi and Karnataka to Maharashtra. In reference to COVID-19, different scenarios of mobility may arise: the mobility can be (a)towards the native site, (b) towards the professional place or (c) nearly equal in both the directions. As an example, few days before the declaration of lockdown, the migrants might prefer to move towards their native places; on the closing of the lockdown period, the migrants may flow back to their professional sites and on the usual days the transport can be nearly equal in both the directions. The color map of figures 1b and 1c are depicting the total number of reported cases normalized with per million population. Throughout this article, we have considered the cases reported on April 10, 2020, and one can easily replace it with current data before the analysis and migration factor can also be reduced based on the situation. The case density (in per million population) is highly diverse, with a maximum at Delhi (40), Andaman and Nicobar Islands(29), Chandigarh(17), Jammu, Kashmir and Ladakh(14), Kerala(10), Tamil Nadu(10)and Maharashtra(10). Figure 1b represents the flow of people towards their native places (case a), which is a question of national concern as tweeted by Environment and Tourism Minister Aaditya Thackeray “Right from the day the trains have been shut down, the State had requested trains to run for 24 hours more so that migrant labor could go back home. CM Uddhav Thackeray ji raised this issue in the PM-CM Video Conf as well as requesting a roadmap for migrant labor to reach home.” However, the preliminary calculations (Figure 1b and 1d) reveal that this transport may transfer a significant number of infected individuals to Uttar Pradesh and Bihar and can multiply further in the packed trains. It may be noted that, if the same scenario would occur in any other state having lower cases, instead of Maharashtra, then the problem might not be that much challenging. Now, we study case (b), i.e. the flow of people is from native to professional sites, which usually occurs at the ending of lockdown. Figure 1c and 1e show that Maharastra, Karnataka, Haryana, Uttar Pradesh could receive maximum infected individuals. Strikingly, Karnataka, Haryana and Uttar Pradesh were not having high existing cases, yet could receive significant cases during the ending of lockdown. Further, Haryana and Uttar Pradesh may receive high cases from the hotspot Delhi, while Karnataka may receive infected cases from Andhra Pradesh, Telangana, Kerala, and Tamil Nadu. On the other hand, Kerala, which emerged as one of the first hotspots, with several international cases, may not receive significant national cases due to fewer migrants. Now we study case(c), importantly, it represents the natural connectedness between two locations, and in turn, decides the regular human mobility.. After a certain interval from lockdown, the flow would become equal from both sides due to regular movements; the transport, in this case, is the average of both side migrant flow (Figure 1a). The analysis of regular flow shows that Uttar Pradesh, Bihar, Maharastra, and Karnataka can be at more risk, once the transport is resumed. By the above discussion, we conclude that the spreading depends on existing cases, connectivity between locations and social scenario, and is somewhat predictable by the proposed model.

**FIG. 1.**
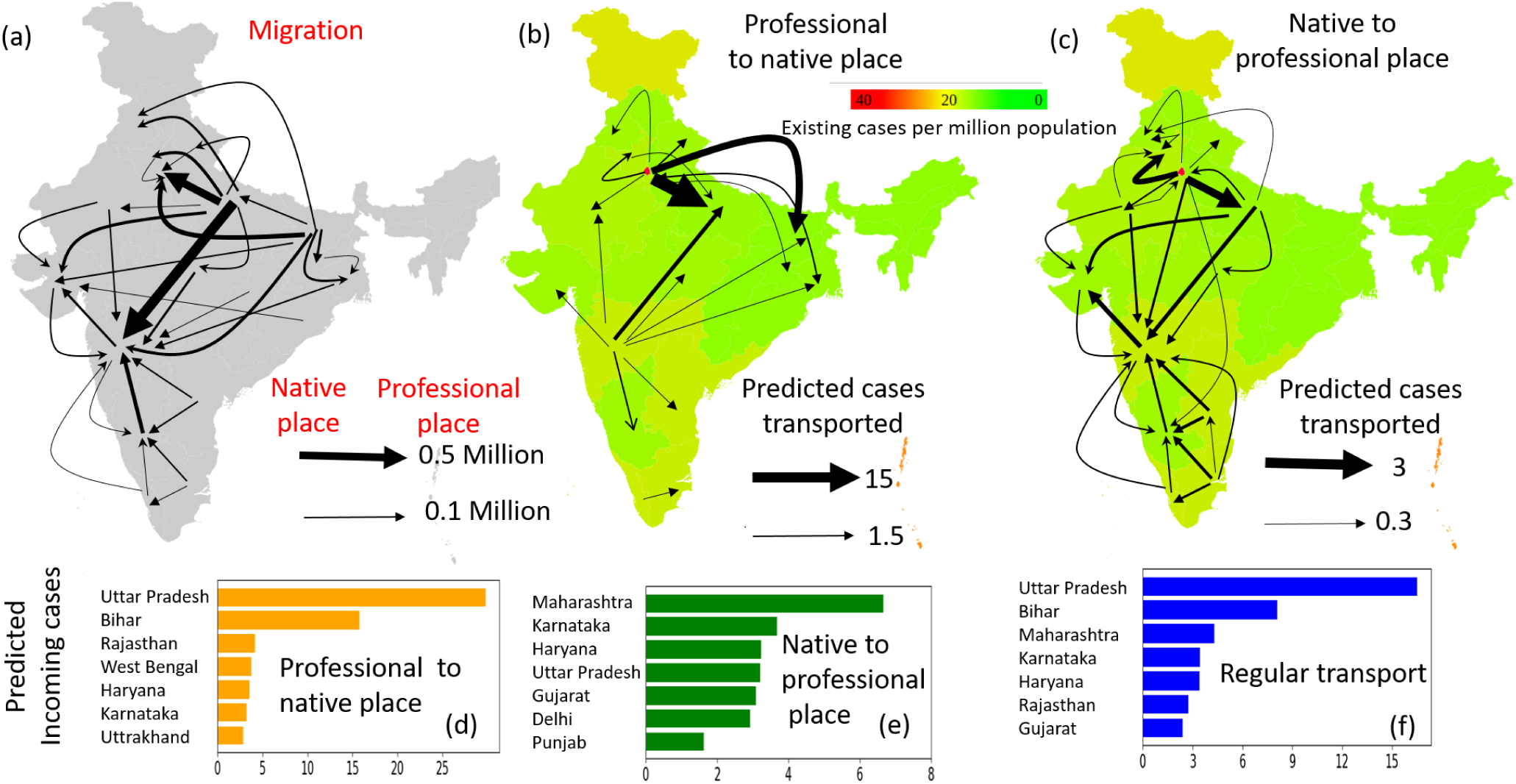
(a)Arrow map showing migration profile between different states. The direction of arrow is from native to professional place, and the arrow width represents the migration population. (b-c) Color map of reported COVID-19 cases per million population (as per April, 10, 2020). (b) The arrows represent the esitamated number of infected individuals from professional to native places and (c) native to professional place. List of states with maximum incoming infected individuals during (d) professional to native place transport, (e) native place to professional place transport and (f) during regular transport.

Next, we examine the spread of COVID-19 via transport through the airline’s network. To obtain the mobility of passengers via airflow, we use monthly airline data provided by the Ministry of Civil Aviation.[20] Figure 2a represents the pairwise passenger flow among different airports. The predicted flow of infected passengers is calculated and shown in figure 2b. It is clear from figure 2b, the cities with passengers from hotspots like Delhi, Mumbai can receive significant cases. Figure 2c shows the predicted number of incoming infected passengers at 40 busiest airports. Mumbai, Delhi, and Bangalore would be receiving maximum incoming infected individuals, which is expected because of their high traffic. To normalize this effect, the total number of infected passengers is divided by the total number of passengers. The airports Amritsar, Dehradun, Srinagar, Jodhpur, Lucknow, Jammu, Patna, Varanasi can significantly receive a higher number of normalized infected individuals; it may be noted that all of these airports are situated in north India. Also, it is interesting to know that most of the North-East airports (Imphal, Agartala, and Guwhati) and a few south Indian airports (Vishakhapatnam and Madurai) are relatively at the lower level of danger, due to less connection with hotspots like Delhi and Mumbai. The work can help in identifying which airports could be shut down, which could remain operational and estimating the required number of quarantine and medical facilities.

**FIG. 2.**
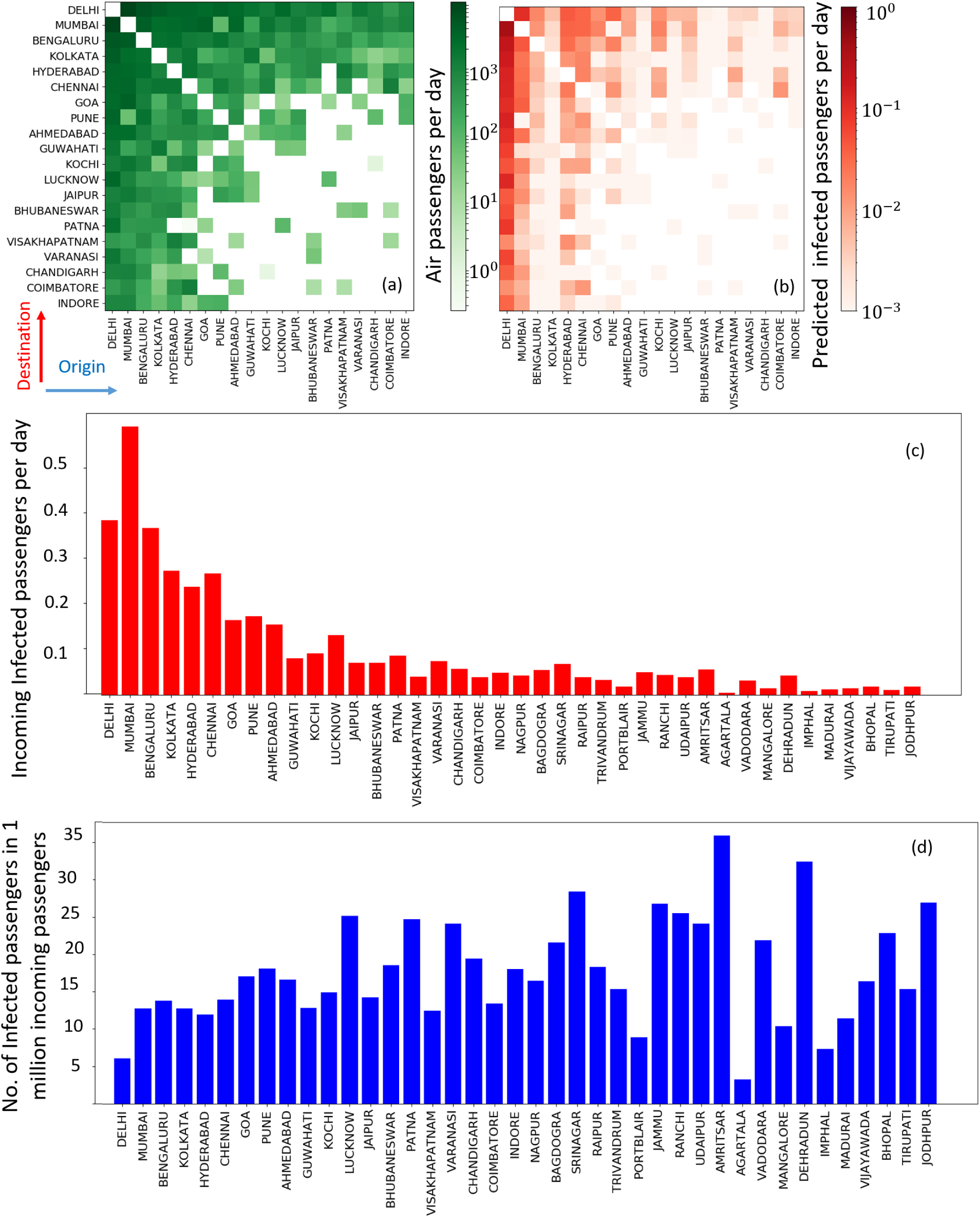
(a) Pairwise number of passengers traveling between various airports. Only data of 20 airports (out of 75) are shown for visualization. (b) The predicted number of infected passengers traveling between different airports. (c) The predicted number of total incoming infected passengers at different airports (d) and normalized incoming infected passengers at different airports.

Through the above discussion, we infer that the incoming cases based on national migration cases are not going well with the current cases. The passenger’s air-transport analysis and regular transport based on migration map (case(c)), both predict that Uttar Pradesh, Bihar, and Karnataka could receive a significantly higher number of cases. Maharashtra with higher current cases already can receive additional incoming cases. Several unpredictable local factors and strictness of administration may affect the active cases dynamically, thus the analysis should be performed on a day to day basis, once the transport is resumed. Note that the airport traffic flow is an indicator of connectedness between two locations, thus results by analyzing train or road network may also provide similar inferences. The developed model and platform can effectively be used for real-time risk monitoring of different locations. Therefore, future research will give optimal techniques for managing airflow, with minimum case-spread and maximum airflow. The model can also be used for border policy purposes, in an international situation that differs dynamically. For instance, hotspots shifted dramatically within the first four months of 2020, hotspots began from China in January, impacted Iran in February, Italy and Europe in March, and the US in April. Not that, India’s foreign mobility is very complex from one state to another. As a result, higher cases of COVID-19 in UAE can have a greater impact on Kerala, whereas the higher cases in Canada can have a greater impact on Punjab. The model may provide a systematic approach for controlling international flights in a rapidly evolving COVID chart. The model is generic and has the potential to study more magnified regions such as state or districts; mobile location data, bus, or train passenger’s network data can be used for the purpose. Future work can investigate the time evolution aspect of COVID-19 in different locations.

## III. CONCLUSION

The present work proposes a network-based model for predicting the spread of COVID-19, incorporating human mobility through knowledge on migration and air-transport. We found that migration towards native places may result in higher incoming cases in Uttar Pradesh and Bihar, while migration to the professional sites can surge incoming cases in Karnataka, Maharashtra, and Haryana, and daily flows are likely to endanger both sets of sites. Airports situated in North India are relatively at higher risks as compared to northeast and south. The model could be utilized to estimate the number of quarantine and medical facilities for incoming cases. Additionally, it will help to control the spread of COVID-19 by closing specific routes with a higher risk of pandemic spread.

## Data Availability

Not applicable.

## ABOUT THE AUTHOR

Ankush Kumar received the M.S.-Ph.D. degree from the Jawaharlal Nehru Centre for Advanced Scientific Research, Bengaluru, in 2018, under the supervision of Prof. G. U. Kulkarni. He completed his first Post-Doctoral at Department of Mathematics, University of Pittsburgh and currently a Post-Doctoral Associate at Institut d’Électronique de Microélectronique et de Nanotechnologie, France. His research interests are modeling on networks, devices, and neuromorphic systems.

